# Spectral imaging enables contrast agent-free real-time ischemia monitoring in laparoscopic surgery

**DOI:** 10.1101/2022.03.08.22271465

**Authors:** Leonardo Ayala, Tim J. Adler, Silvia Seidlitz, Sebastian Wirkert, Christina Engels, Alexander Seitel, Jan Sellner, Alexey Aksenov, Matthias Bodenbach, Pia Bader, Sebastian Baron, Anant Vemuri, Manuel Wiesenfarth, Nicholas Schreck, Diana Mindroc, Minu Tizabi, Sebastian Pirmann, Brittaney Everitt, Annette Kopp-Schneider, Dogu Teber, Lena Maier-Hein

## Abstract

Laparoscopic surgery has evolved as a key technique for cancer diagnosis and therapy. While characterization of the tissue perfusion is crucial in various procedures, such as partial nephrectomy, doing so by means of visual inspection remains highly challenging. Spectral imaging takes advantage of the fact that different tissue components have unique optical properties to recover relevant information on tissue function such as ischemia. However, clinical success stories for advancing laparoscopic surgery with spectral imaging are lacking to date. To address this bottleneck, we developed the first laparoscopic real-time multispectral imaging (MSI) system featuring a compact and lightweight multispectral camera and the possibility to complement the conventional RGB (Red, Green, and Blue) surgical view of the patient with functional information at a video rate of 25 Hz. To account for the high inter-patient variability of human tissue, we phrase the problem of ischemia detection as an out-of-distribution (OoD) detection problem that does not rely on data from any other patient. Using an ensemble of invertible neural networks (INNs) as a core component, our algorithm computes the likelihood of ischemia based on a short (several seconds) video sequence acquired at the beginning of each surgery. A first-in-human trial performed on 10 patients undergoing partial nephrectomy demonstrates the feasibility of our approach for fully-automatic live ischemia monitoring during laparoscopic surgery. Compared to the clinical state-of-the-art approach based on indocyanine green (ICG) fluorescence, the proposed MSI-based method does not require the injection of a contrast agent and is repeatable if the wrong segment has been clamped. Spectral imaging combined with advanced deep learning-based analysis tools could thus evolve as an important tool for fast, efficient, reliable and safe functional imaging in minimally invasive surgery.

## 1 Main

Replacing traditional open surgery with minimally invasive techniques for complicated interventions such as tumor resection is one of the most important challenges in modern healthcare. Conventionally used RGB (Red, Green, and Blue) camera-based laparoscopes, however, are ill-suited for these demands. Their mode of operation is based on mimicking the human eye by collecting light in the aforementioned three broad regions of the optical spectrum; as a consequence, precise tissue differentiation and assessment of organ function remain largely inaccessible. Yet, obtaining real-time functional tissue information is crucial for many key procedures in minimally invasive surgery.

A common necessity of laparoscopic surgeries, for example, is stopping the blood flow to a specific organ or tissue region by clamping the arteries responsible for blood supply. This process, commonly referred to as *ischemia induction*, prevents excessive bleeding of patients^1^ and is performed in various procedures, including partial nephrectomy, organ transplantation and anastomosis. After clamping the main arteries, it is highly challenging to assess the perfusion state of the tissue solely based on the available RGB video stream. This especially holds true for selective clamping of a segmental artery, in which ischemia is induced only in the cancerous part of the kidney during partial nephrectomy^2, 3^. The most common approach to ensure correct clamping is based on indocyanine green (ICG) fluorescence (see Fig. 1): After ICG is injected into the bloodstream, it binds to plasma proteins. The bound ICG travels through the bloodstream and accumulates in the internal organs, especially in the kidney and liver, within a minute^4, 5^. Lack of a fluorescent signal thus corresponds to lack of perfusion. However, due to long washout periods of about 30 minutes, this test is not easily repeatable if the wrong segment has been clamped^5^ or if the clamping procedure was done improperly, as illustrated in Fig. 1. Furthermore, it requires a contrast agent to be injected into the bloodstream. Even though ICG injection is generally regarded as a safe procedure, cases with severe complications such as anaphylactic shock have been observed^6^.

**Figure 1.**
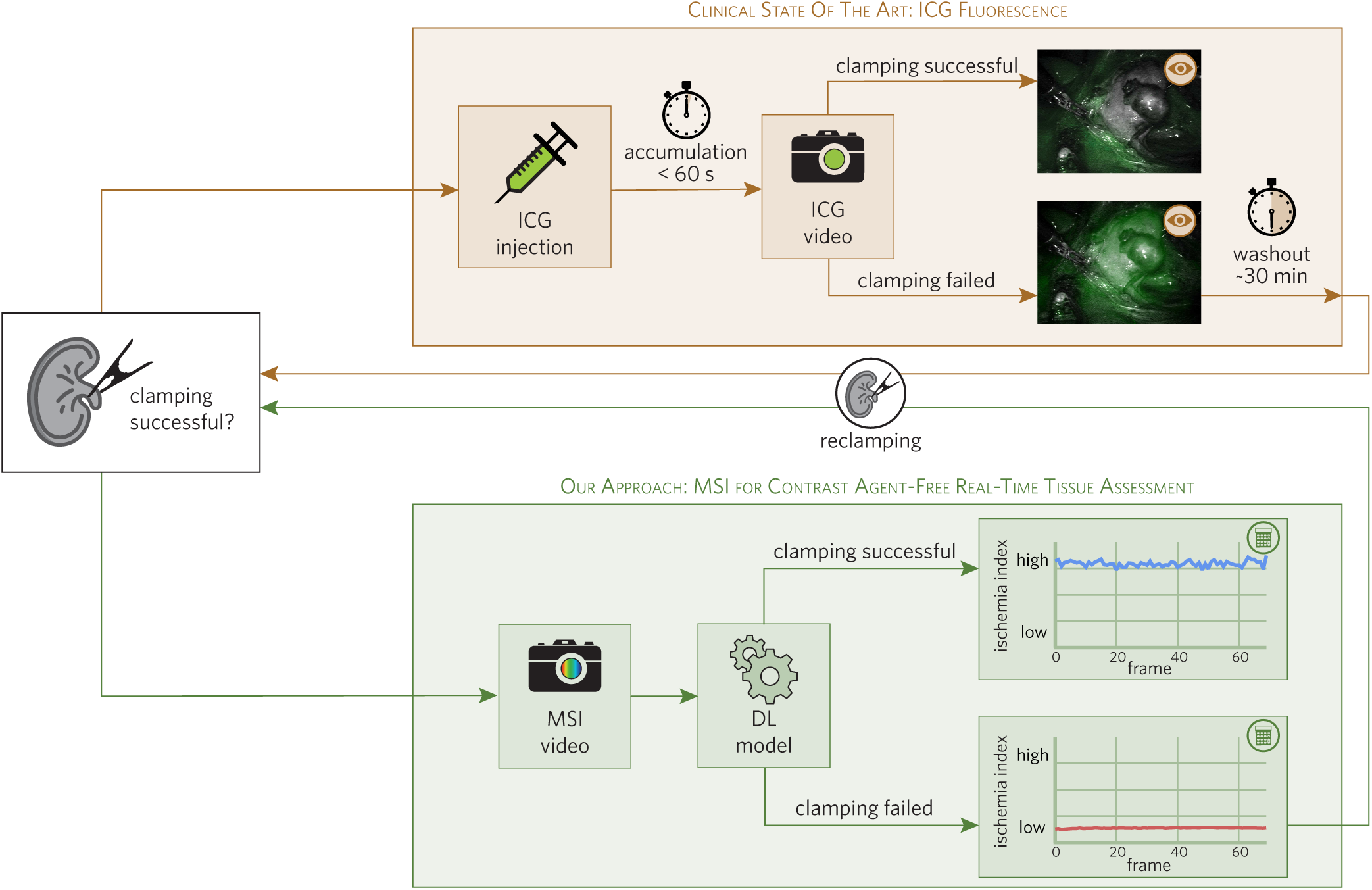
Our multispectral imaging (MSI)-based approach enables continuous, contrast agent-free, real-time ischemia monitoring in laparoscopic surgery. The clinical state of the art (brown) to verify successful ischemia induction via clamping of arteries is based on indocyanine green (ICG) imaging: ischemic tissue is characterized by a lack of fluorescent signal, whereas perfused tissue fluoresces. In the case of unsuccessful clamping, the test can only be repeated after a washout period of about 30 min. Our approach (green) is based on non-invasive and contrast agent-free MSI at video rate. Deep learning (DL) models trained on MSI video sequences prior to clamping are capable of detecting ischemic tissue areas as outliers in real-time, as detailed in Fig. 3.

Spectral imaging is a promising alternative approach to improving surgical vision. This technique removes the arbitrary restriction of recording only three broad spectral bands (Red, Green and Blue) by capturing an *n*-dimensional feature vector for each camera pixel, where each dimension corresponds to a comparatively narrower spectral band. As different tissue structures possess unique optical scattering and absorption properties, knowledge of these optical properties along with spectral measurement data can potentially provide important information on tissue morphology^7–10^ and function^11, 12^. The term multispectral imaging (MSI) is used when only a few bands (up to dozens) are recorded, while hyperspectral imaging (HSI) refers to hundreds of bands being recorded^13^.

Despite the general success of MSI and HSI^13–32^, applications in the operating room (OR) have been limited. Some of the main reasons why spectral imaging has not yet found its way into surgical practice are related to image acquisition time, processing time and size of the available devices^13^. In fact, many available MSI/HSI camera systems are large (14–50 cm) and/or take several seconds (2–8 s) to record and process one image^32–39^. To the best of our knowledge, the only laparoscopic spectral device proposed for clinical use so far^34^ takes around five seconds to record one hyperspectral image, which prevents real-time application. In consequence, clinical success stories in spectral imaging for minimally invasive surgery are still lacking. Specifically, we are not aware of any clinical study in the broader context of real-time perfusion monitoring based on spectral imaging in laparoscopic surgery.

We address this gap in the literature with the following contributions:

1. **Video-rate MSI system** (see Fig. 2): We present the first real-time (25 Hz) laparoscopic MSI system applied in patient studies. It features a compact (26 × 26 × 31 mm) and lightweight (32 g) MSI camera that can be connected to standard laparoscopes via a widely used C-Mount adapter and operates with clinical light sources.

**Figure 2.**
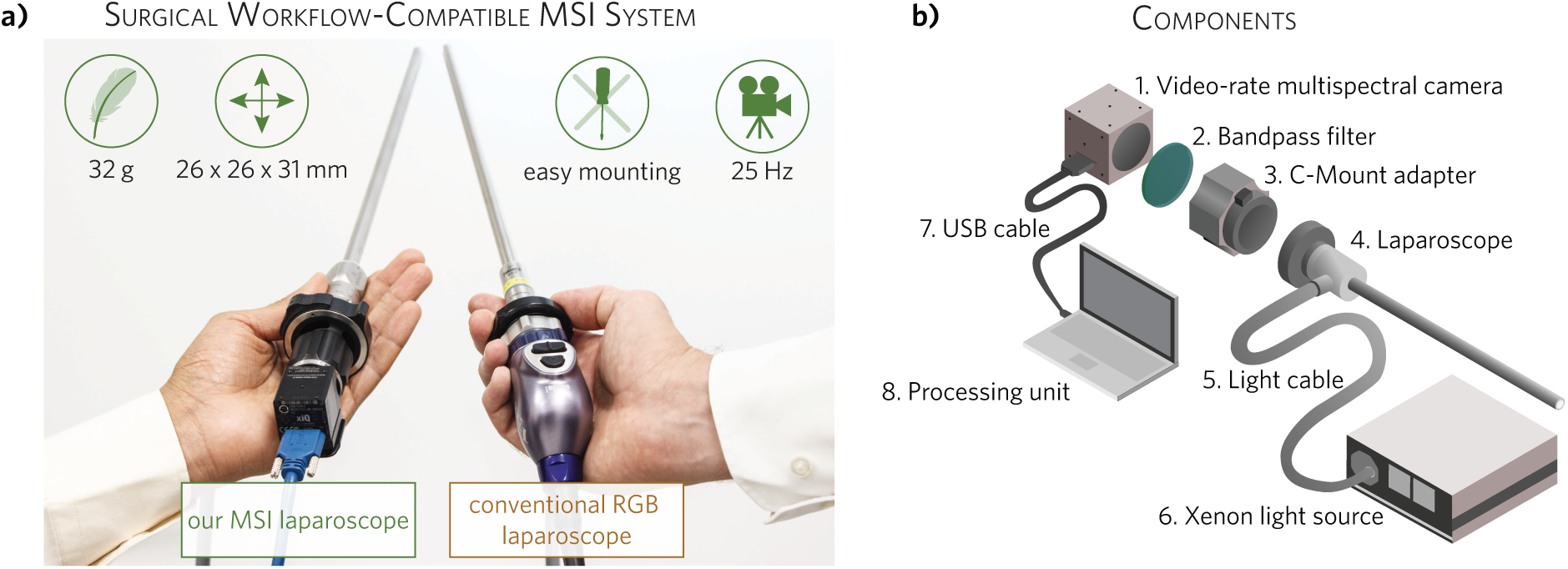
Our multispectral imaging (MSI) system for ischemia monitoring is comparable to current standard equipment in terms of size and weight. **a)** Proposed spectral imaging-based laparoscope (left) compared to standard Red, Green, and Blue (RGB) laparoscope (right). **b)** Schematic representation of the system developed for spectral tissue analysis in laparoscopic surgery (see Sec. 5.1). The dimensions of the laptop and the light source have been scaled down for visualization purposes.
2. **Deep learning-based algorithm for real-time ischemia detection** (see Fig. 3): To overcome the need of large amounts of training data required by traditional discriminative machine learning methods, we phrase the problem of ischemia detection as an out-of-distribution (OoD) detection problem that does not rely on data from any other patient (patent pending). Using an ensemble of invertible neural networks (INNs) as a core component, our algorithm is trained to compute the likelihood of ischemia based on a short (several seconds) video sequence acquired at the beginning of each surgery.

**Figure 3.**
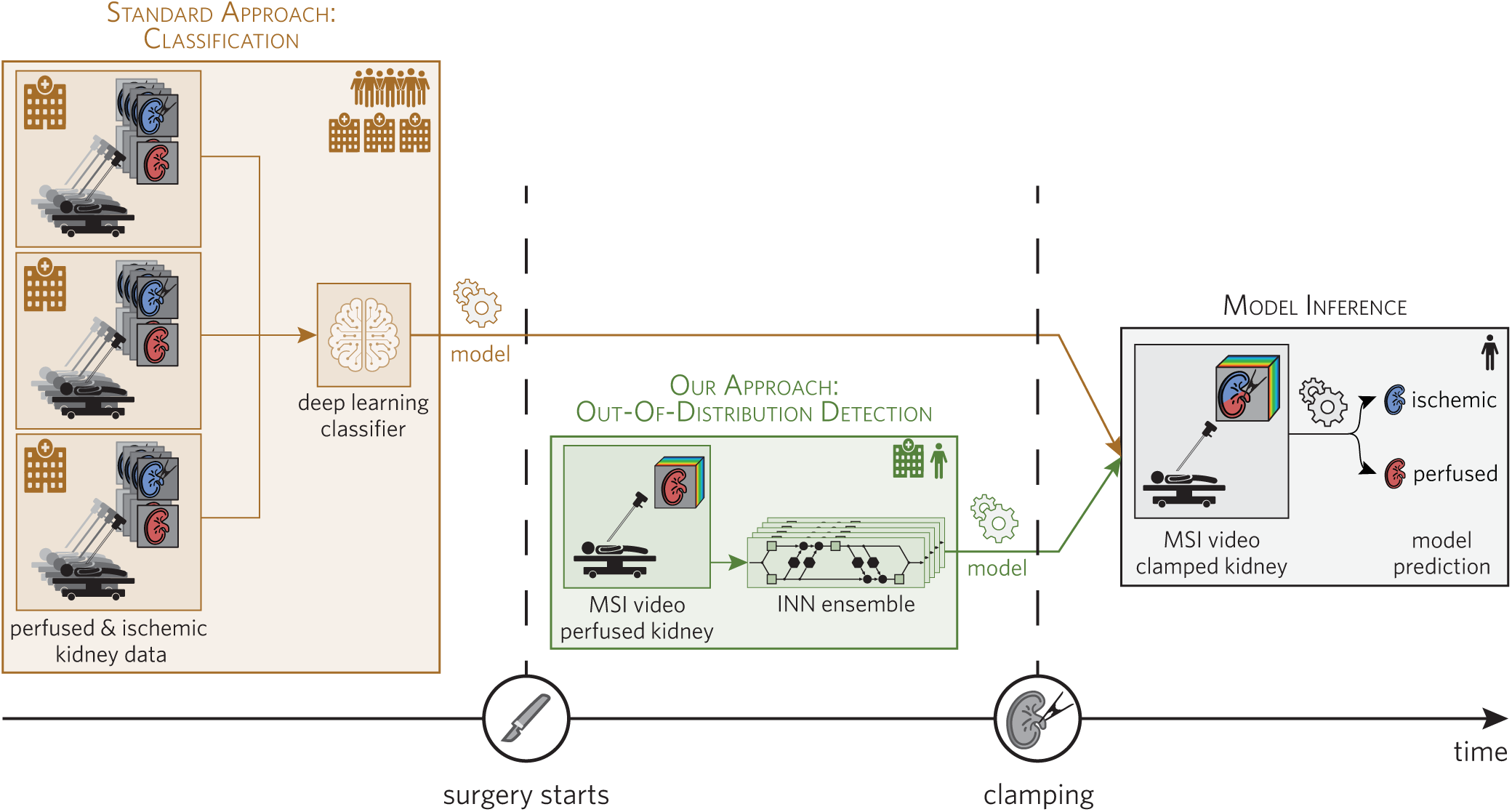
We treat ischemia monitoring based on multispectral imaging (MSI) as an out-of-distribution (OoD) task which only needs training data from one single patient. Traditional deep learning (DL) methods (brown) require large amounts of patient data to train a model, while our method (green) only needs data from a single patient. Using an ensemble of invertible neural networks (INNs) as a core component, our algorithm computes the likelihood of ischemia based on a short (several seconds) video sequence acquired at the beginning of each surgery. An important feature of our approach is that the entire training and inference process can be performed during a surgical procedure.
3. **First-in-human study:** We present the first-in-human study demonstrating that monitoring kidney ischemia in real time is now possible.

## 2 Results

### First-in-human application of video-rate multispectral laparoscope

The proposed system enables us to pioneer clinical video-rate MSI in laparoscopy. As shown in Fig. 2, it comprises (1) a snapshot MSI camera, (2) a 335–610 nm bandpass filter, (3) a C-Mount adapter with adjustable focal length, (4) a standard surgical laparoscope, (5) a surgical light cable, (6) a Xenon light source, and (7) a USB cable to connect the MSI camera to the (8) processing unit (see Sec. 5.1). Due to the compact (26 × 26 × 31 mm) design, the camera does not add hardware complexity to the OR (see Fig. 2 a). Furthermore, the camera is extremely lightweight (32 g), thus enabling easy handling of the device over long periods of time. The system operates at a frame rate of 25 Hz. To compensate for tissue and camera motion during image acquisition, we developed a deep learning-based algorithm capable of tracking regions of interest (ROIs) within an MSI video (see Sec. 5.4). The algorithm was successfully applied in 10 patients undergoing partial nephrectomy and served as a prerequisite for further analysis and for computing the deep learning-based *ischemia index*. The results of our analysis are exemplary illustrated in this video.

### High inter-patient variability for kidney tissue

High inter-patient variability generally suggests poor generalizability of supervised learning algorithms. In a recent porcine study we showed that the greatest source of variability related to spectral images of organs acquired from healthy animals is the organ under observation rather than the recorded individual or specific acquisition conditions^40^. This enabled us to develop a highly accurate supervised deep learning (DL) algorithm for fully-automatic organ classification^41^. However, an analogous analysis applied to the data of the clinical study presented here revealed that kidney tissue of patients undergoing partial nephrectomy is highly heterogeneous. As illustrated in Fig. 4, when reducing the full spectral information to two dimensions via principal component analysis (PCA)^42^, the measurements from different patients gather within clear visual clusters for each state (indicated by a star (perfused) or circle (ischemic) in Fig. 4 a), while a clustering of different tissue states across patients cannot be observed (circles and stars do not form clear clusters). Note in this context that the first two principal components (PCs) capture 87 % of the variation. Furthermore, according to a mixed model analysis, most variability in the measurements can be explained by the individual patient rather than—as would be desired—the tissue state (see Fig. 4 b)). In consequence, tackling the challenge of ischemia detection with a traditional supervised algorithm trained on a small data set would come with a high risk of poor generalization capabilities to unseen individuals. In addition, (slight) changes in the acquisition setup might require complete retraining of the method on the entire patient database. This motivated our personalized approach to ischemia detection, illustrated in Fig. 3.

**Figure 4.**
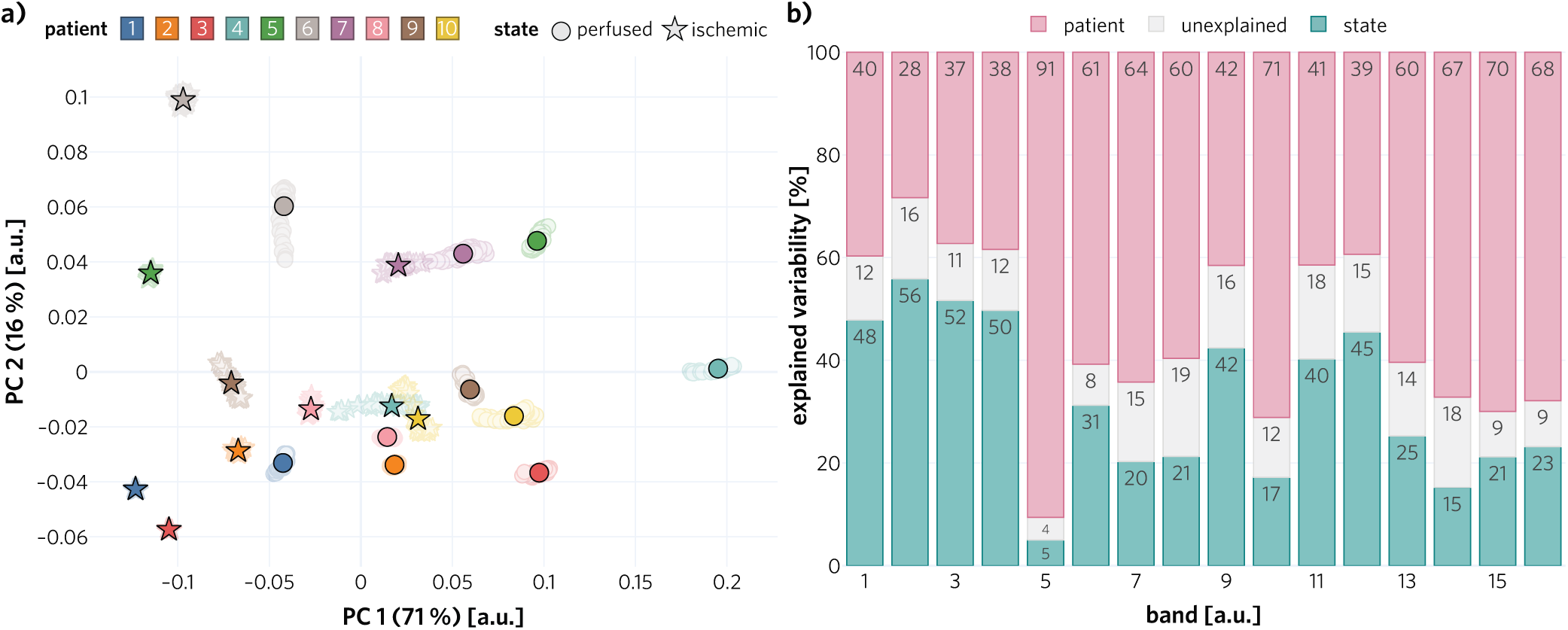
High tissue heterogeneity across subjects motivated our personalized approach. Both the **a)** principal component analysis (PCA) and **b)** the mixed model analysis demonstrate the high inter-patient variability of spectral tissue measurements. **a)** The fact that different tissue states cluster within a subject but do not form a uniform cluster across subjects motivated us to phrase the challenge of ischemia detection as an out-of-distribution (OoD) problem. Solid markers show the cluster centers, transparent markers the raw data points and the axis labels denote the explained variance of the corresponding principal component (PC). **b)** Explained variance for the 16 bands of the multispectral imaging (MSI) camera, depicted in Fig. 7

### Novel deep learning-based *ischemia index* captures ischemia state

To overcome the limitations of supervised approaches in the presence of high tissue heterogeneity, we developed a personalized approach to ischemia detection that solely requires data of the patient undergoing surgery (see Fig. 3). Specifically, we phrase the problem of ischemia detection as an OoD detection problem and assess the perfusion state with a custom-designed *ischemia index* that relies on an ensemble of INNs as the core component. As detailed in Sec. 5.2, the ensemble is trained to estimate the density of the distribution of perfused spectra from a short (several seconds) video sequence acquired at the beginning of each surgery. At inference time, the density is evaluated on new spectra and the ensemble predictions are aggregated using the Widely Applicable Information Criterion (WAIC)^43^. The *ischemia index*, computed for each image, represents the spatially aggregated WAIC values. According to our patient study with 10 patients undergoing partial nephrectomy, in which ICG fluorescence served as the gold standard method, our novel *ischemia index* classifies the perfusion state with high accuracy (see Fig. 5). In fact, in almost all patients, the data corresponding to ischemic tissue could be perfectly separated from those corresponding to perfused tissue. This led to a median / mean area under the receiver operating curve (AU-ROC) of 1.0 / 0.9 obtained for the *n* = 10 patients. While the patient-individual training took 30 s per network on average, the actual index calculation runs at 120 Hz on the selected ROIs on the kidney, and thus in real time.

**Figure 5.**
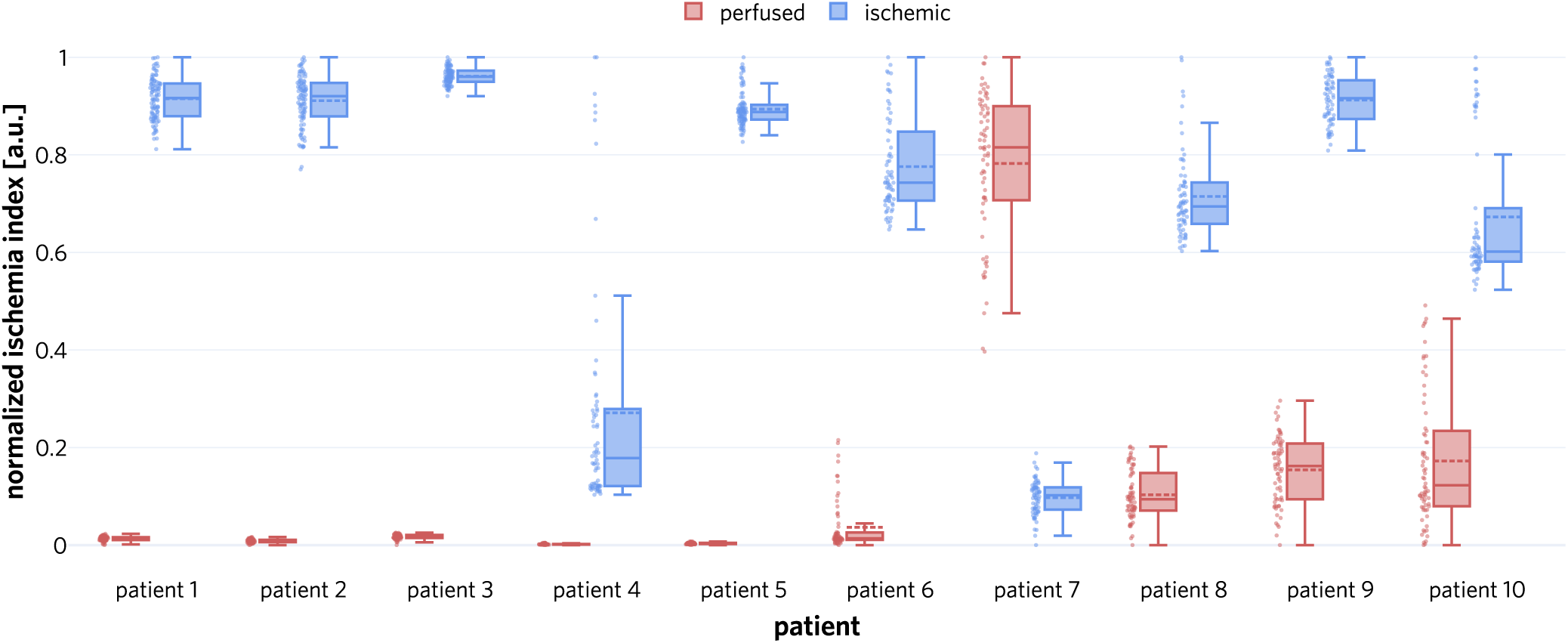
Our approach is capable of discriminating between ischemic and perfused kidney states. We calculated the *ischemia index* for every frame in video sequences of perfused and ischemic kidney separately for each patient (see Sec. 5.2) and generated corresponding dot and box plots. The boxes show the interquartile range with the median (solid line) and mean (dashed line) while the whiskers extend up to 1.5 the interquartile range. Min-max-normalization was performed for clarity of presentation.

## 3 Discussion

MSI is on the verge of opening up entirely new avenues in the early detection, diagnosis and monitoring of diseases ranging from cancer to cardiovascular and inflammatory diseases, particularly when applied in modern interventional healthcare such as minimally invasive surgery. To the best of our knowledge, this work presents the first ever real-time application and analysis of MSI in laparoscopic surgery on human subjects and in a clinical setting. Based on a newly developed video-rate multispectral laparoscopic imaging system, our approach presents three key advantages over the current state of the art in interventional ischemia detection:

1. **No contrast agent:** Our method is completely non-invasive and does not require the administration of ICG or any other contrast agent. This reduces intervention time and costs and enhances patient safety by reducing complication risks (e. g. anaphylactic shock).
2. **Multiple uses per surgery possible:** In case of a failed ischemia induction attempt, our method can be reapplied instantly. There is no washout period.
3. **Video-rate ischemia monitoring:** The proposed *ischemia index* can be computed in real time, making continuous ischemia monitoring feasible, whereas the classical approach allows for only a single measurement per surgery.

In the following sections, we discuss our hardware choices to achieve video-rate ischemia monitoring, the consequences of high tissue heterogeneity, our approach to ischemia detection and a comparison between our proposed approach and current clinical practice.

### 3.1 MSI system design

We developed the first real-time MSI system for laparoscopic surgery that was applied in a patient study. The key strengths of our imaging system are the high acquisition speed (above 25 MSI images per second), the low weight (32 g), the small size (camera cube with edge size of 26 × 26 × 31 mm) and the fact that it operates with a standard clinical light source, all of which allow for smooth integration into the surgical workflow. Furthermore, all relevant changes made compared to the standard clinical setup concern components that are attached to the laparoscope and as such never touch the patient, which minimizes the patient risk introduced by a new imaging device.

Previous work on clinical spectral imaging has focused on systems for open surgery with acquisition times of several seconds^32–39^. Preclinical multispectral or hyperspectral systems proposed in the specific context of laparoscopy have typically been based on sequential image acquisition^11, 44–46^, e.g. using the filter wheel technology^11, 44, 47^, and thus come with the risk of motion artefacts and the lack of video-rate acquisition capabilities. The snapshot technique applied in this work has in parallel been explored by other authors^48^ in the context of open surgery. Overall, we are not aware of any real-time MSI or HSI laparoscopic system that has been applied in humans so far.

While the snapshot technique is the key enabler for fast image recordings, it also comes with several limitations. The second order peaks of the MSI camera filters shown in Fig. 7 can potentially present challenges to image interpretation and analysis. Given the fact that these second order peaks are located at low wavelengths in the optical range, and that oxyhemoglobin (HbO_2_) and deoxyhemoglobin (Hb) absorb light mainly in those spectral regions, they need to be taken into account when the target application is quantitative perfusion or oxygenation estimation. For example, applying the Beer-Lambert law^44, 49^ to our data yielded implausible values (e. g. oxygenation above 100%), as discussed below. Our proposed method is not influenced by these second order peaks because the INNs learn the density of the MSI data, regardless of the influence such peaks have on the reflectance spectra. More specifically, given that all image sequences are equally influenced by these second order peaks, INNs encode such influences in the model and learn to predict our *ischemia index* based only on physiological changes and not on hardware configurations.

At the moment, the camera operates in the visible spectrum. However, the tissue spectrum in the infrared (IR) range is often very expressive^10^, promising even better performance in estimating tissue parameters. It could thus be viable to explore this regime with a combination of cameras receptive in the IR range and IR optimized light sources. Another necessary adaptation of the camera system would be the capability to provide both a regular RGB image for human-interpretable visualization to the surgical staff as well as MSI data for the *ischemia index* computation.

### 3.2 Tissue heterogeneity and personalized approach

To our knowledge, this work constitutes the first systematic *in vivo* spectral analysis of diseased kidney tissue. Our analysis clearly exposed the high inter-patient variability of optical properties. A key finding was that the main contributor to the variance of spectral measurements is the patient rather than the perfusion state of the kidney. These results dim the hopes of successfully applying supervised machine learning algorithms as ischemia detectors under conditions of limited patient data.

Our findings regarding the heterogeneity are in stark contrast to recent findings in porcine organs^40^, where the influence of the different specimen is small compared to other explanatory variables. A key difference between this study and the present work (apart from the species) is that for the porcine study all organs were healthy, whereas all of our human subjects underwent partial nephrectomy due to kidney cancer. This might be one factor explaining the high inter-patient variability.

The limited number of patients included in our study makes it hard to determine the mechanism for the change in spectra. In particular, the combination of cancer type and comorbidities is unique for each patient. Furthermore, the complex environment of the OR offers a significant number of external factors that cannot be controlled. For example, the pose of the laparoscope relative to the kidney changes between each patient due to variables such as port placement, respiratory movements, different anatomy, etc. Since the laparoscope is operated freehand, additional motion from the surgeon is involved. Data normalization can only compensate for some of these factors. Overall, the described mismatch between sample size and the number of variable factors makes attribution of spectral variability to these factors hard. However, they are captured as the remaining unexplained variance in Fig. 4 b).

We observed that the explained variability of bands 5 and 10 that can be accounted for by changes in tissue state (perfused *vs*. ischemic) is especially low in comparison to the variability explained by different patients (see Fig. 4). We attribute this to two main reasons: a) The maxima of the camera filter responses corresponding to these bands are located at wavelengths where the extinction coefficient of Hb and HbO_2_ are very similar (see Fig. 7). This, in turn, leads to lower variability between tissue states. b) Band 5 is also affected by a second order peak with a maximum filter response located at a wavelength where the extinction coefficients of Hb and HbO_2_ are considerably smaller, thus leading to higher reflectances and a lower influence of different tissue states (perfused *vs*. ischemic) in the reflectance signal.

In the future, the incorporation of more explanatory variables (such as comorbidities) in our linear mixed model would allow for an in-depth understanding of how different factors impact spectral measurements. However, with the frequency of partial nephrectomies being limited, particularly in pandemic times, additional data acquisition is a slow process. Obtaining an MSI imaging device certified for clinical use can be considered a pivotal step in accelerating data acquisition for large-scale studies.

### 3.3 Ischemia index

We introduced an entirely novel approach to ischemia detection which neither relies on potentially oversimplified modeling assumptions (such as the Beer-Lambert law) nor on a large patient cohort for algorithm training. Our results show that we can overcome difficulties posed by the high inter-patient variability through rephrasing the problem of ischemia detection as an OoD detection problem. In the vein of personalized medicine, this process is solely dependent on data from the patient in question, which avoids the necessity for consideration of confounders introduced by different patients^33, 50^. This constitutes an extremely relevant feature in the face of increasing evidence that many research studies overestimate the performance of DL algorithms due to poor selection of test data relative to the training data^51, 52^. Furthermore, algorithms trained on data from a specific camera may not necessarily generalize to slightly adapted conditions^33, 53^. Even though our model has to be retrained for every patient, application in the OR is feasible as only spectra of perfused tissue are required for training and the training time is approximately 30 s per network (five networks). Such a training procedure can easily be performed at the beginning of each surgery prior to identification of the renal artery, thus avoiding delays in the surgical procedure. Furthermore, at a spectrum evaluation rate of 220 kHz, which translates to an individual ROI evaluation rate of 240 Hz, the inference is real-time capable. The clinical state-of-the-art method for assessing ischemia in partial nephrectomy is based on ICG fluorescence. As summarized in Tab. 1, key advantages of our method are its non-invasiveness and real-time capability. A limitation of our approach can be seen in the fact that our algorithm requires a clean kidney to perform reliably. Excessive bleeding, scarring, or remaining fatty tissue on the kidney surface in particular may hinder the application of our method. On the other hand, this disadvantage may potentially be overcome by extending the wavelength range to the IR range, which is associated with deeper tissue penetration.

**Table 1.** Key characteristics of traditional ischemia monitoring with ICG injection and our proposed non-invasive method.

| Feature | ICG | Our method |
| --- | --- | --- |
| Standard light source | X | ✓ |
| Standard camera | X | X |
| Low per use cost | X | ✓ |
| Low investment cost | X | X |
| Contrast agent-free | X | ✓ |
| Multiple use per surgery | X | ✓ |
| Robust to tissue surface imperfections | ✓ | X |
| Real-time | X | ✓ |

The state-of-the-art approach to assessing perfusion based on spectral measurements in preclinical studies is to apply the Beer-Lambert law^44, 49^ (see Sec. 5.4). However, this yielded implausible values (e. g. oxygenation above 100%). In addition, a clear separation of perfusion states could only be achieved in 50% / 30% of the patients, depending on whether oxygenation or blood volume fraction estimation was used as a decision score.

Other model-based approaches to estimating physiological parameters, such as the one applied by the TIVITA^®^ cameras (Diaspective Vision GmbH, Am Salzhaff-Pepelow, Germany), rely on first and second order derivatives of the spectra^46^. Such an approach thus requires fine-grained spectra and is potentially highly sensitive to noise. In consequence, its application has so far been restricted to HSI camera setups associated with long acquisition times.

In our own previous work, we presented a machine learning-based approach to physiological parameter quantification based on MSI data^11, 12^. To address the absence of a reliable reference method for generating labeled training data, we based our approach on simulations generated with Monte Carlo (MC) methods. To this end, we leveraged prior knowledge on tissue composition, optical properties and light-tissue-interaction to generate a large pool of synthetic spectra, labeled with corresponding ground truth physiological parameters. This data was then used to train a machine learning-based regressor^11, 12, 54^, possibly after a domain adaptation step^12^. We did not apply the method in this study as our kidney measurements were OoD compared to the simulations, which indicates that more work is needed for fully realistic spectra generation in the presence of pathologies. To overcome the lack of accurate prior knowledge as required by model-based approaches or the simulation-based model, we decided to explore the personalized method presented in this manuscript.

While our method worked perfectly on nine out of ten patients, it failed on patient 7. Further analysis revealed that the spectra between different kidney states did not substantially differ for this particular patient, as shown in Fig. 6. In fact, a kernel density estimation (KDE) performed on the data resulting from a patient-specific PCA showed a considerable overlap between tissue states. Furthermore, the perfused spectra exhibited higher variability than the ischemic spectra consistently across all bands. These two factors might explain why ischemic spectra were erroneously detected as *in domain*, with the support of the distribution of perfused spectra containing the support of the distribution of ischemic spectra. A possible reason for the similar spectra in perfused and ischemic state is the appearance of burned fatty tissue in the surface of the kidney, which can occur while removing fatty tissue with the da Vinci^®^ monopolar scissors and negatively impact tissue penetration. To mitigate problems related to tissue penetration and thus preclude failure of our method, we envision increasing the wavelength range of the camera and the light source in the future, as mentioned above. Since this change requires a new hardware setup (higher power light source) with regulatory approval, it remained beyond the scope of our current study. It should also be mentioned that the laparoscopic videos of patient 7 looked unusual from a clinical perspective (see Fig. 6 a)). As patient 7 also happened to be the only smoker among the participants of our study, we cannot rule out a possible effect on the measured spectra. Investigating the impact of comorbidities on MSI-based ischemia detection and potential failure cases was not within the scope of the present work, but could be the subject of a more comprehensive study in the future.

**Figure 6.**
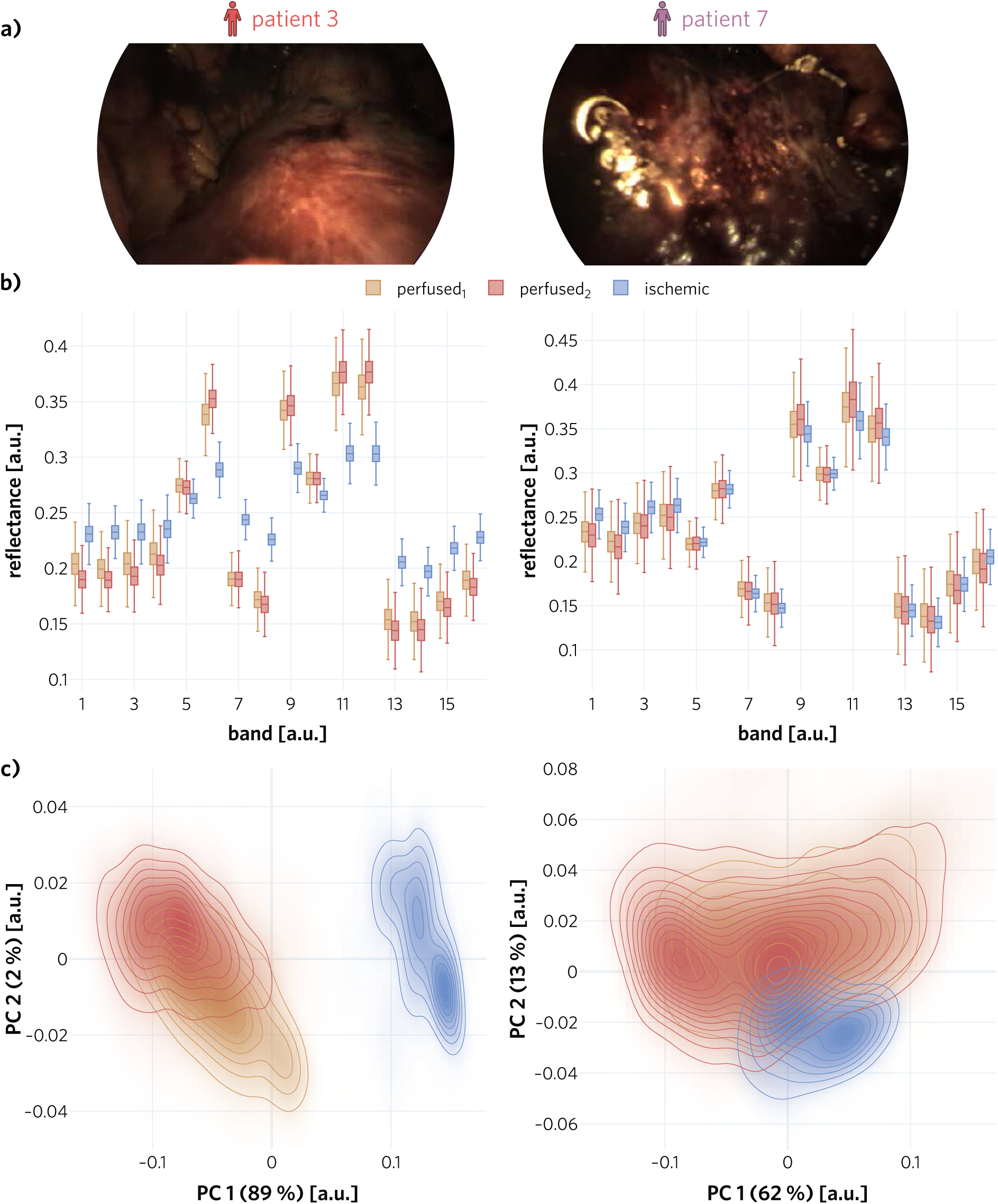
Comparison of a representative patient with patient 7 with respect to the algorithm input. **a)** Example Red, Green and Blue (RGB) images were selected from the perfused_2_ sequence to illustrate the unusual images acquired of the kidney of patient 7. **b)** The reflectances of the representative patient (patient 3) differ clearly between perfused and ischemic tissue for the vast majority of camera bands, while no clear separation can be observed in patient 7. The boxes show the interquartile range with the median (solid line) while the whiskers extend up to 1.5 the interquartile range. **c)** A kernel density estimation (KDE) performed on the data resulting from a patient-specific principal component analysis (PCA) provides further explanation for why our method falsely detected the ischemic kidney data of patient 7 as inlier. The axes labels denote the explained variance of the corresponding principal component (PC).

Of note, patient 4 presented a particular case in our analysis. Due to data corruption of the white Spectralon^®^ reference measurements (see Sec. 5.3.2), we used the white measurements recorded during another surgery with identical hardware setup (patient 3). Given that hardware, software and Spectralon^®^ target were all identical, we did not expect a major influence of this procedure in the analysis of the data from patient 4. Indeed, experiments where the data from patient 4 was normalized using the white and dark references from other patients revealed that the variation across different normalization strategies is insignificant compared to the variance across kidney states (perfused and ischemic).

A specific challenge within this study was the fact that we needed to remove our laparoscope between training and testing data acquisitions. The reason was that we did not want to alter the clinical procedure more than necessary and thus based the clinical decision making exclusively on a standard (additional) RGB laparoscope. In order not to require an additional port for our own laparoscope (which would have come at the cost of increased invasiveness), we used the port that is required for the clamping for our device. This made temporary retraction necessary, thus complicating the ischemia detection because analysis on the same ROI could not be guaranteed. Note that in some of the patients, it was not even manually possible to robustly locate the same tissue region before and after clamping due to the lack of landmarks and the altered pose of the laparoscope relative to the tissue. To ensure that our OoD detector does not merely detect different tissue regions (rather than tissue states), we took the design decision to base the analysis on two disjoint ROIs. Furthermore, we designed the validation study in a way that we acquired data from perfused and ischemic tissue in a comparable manner; always after removal and re-insertion of the laparoscope. If the reason for OoD detection in the ischemic tissue was the change of tissue region, the algorithm would have failed on the testing sequences corresponding to perfused tissue (perfused_2_). Of note, the challenge of interrupted image acquisition may easily be overcome in the future with a clinically certified MSI laparoscope, enabling MSI acquisition and physician interpretation at the same time.

It is further worth mentioning that we investigated applying our *ischemia index* to (reconstructed) RGB data. While a separation of perfusion states was possible for some of the patients, the MSI results were far better in terms of data separation. Working on an even broader wavelength range that incorporates IR bands may improve the results further.

In conclusion, we presented the first approach to ischemia detection based on MSI data in laparoscopic surgery that was successfully applied in a clinical study. Future work should be directed to large cohort studies and the transformation of our index into an easy-to-use score with inherent classification and uncertainty quantification capabilities.

## 4 Conclusion

Here, we presented the first-in-human study demonstrating that intraoperatively monitoring kidney perfusion in real time is possible by exploiting a combination of spectral imaging and advanced deep learning-based analysis algorithms. Our study can be seen as a first foray into the application of multispectral imaging in a clinical interventional setting.

## 5 Methods

This section presents the methods corresponding to the three primary contributions of our paper, namely, the MSI system (see Sec. 5.1), the algorithm for ischemia detection (see Sec. 5.2) and our clinical study for validating our approach (see Sec. 5.3). Implementation details on some of the modules and methods have been moved to a separate section (see Sec. 5.4).

### 5.1 Multispectral imaging system

Our MSI system is illustrated in Fig. 2 and comprises the following main components.

#### Multispectral camera

The core component is an MSI camera (MQ022HG-IM-SM4x4-VIS, XIMEA GmbH, Münster, Germany), which is a small (26 × 26 × 31 mm) and light (32 g). It is based on the imec (Leuven, Belgium) mosaic snapshot sensor, which acquires 16 spectral bands at a single snapshot, using a 4 × 4 repeating mosaic pattern of filters (see Fig. 7 b)); the spectral response of each filter is shown in Fig. 7 c). Several of the bands show two peaks in the spectral response. These are caused by the measurement principle of the Fabry-Pérot filters^55^, which lead to so-called “second order” peaks. The intensity of such peaks depends on the height of the optical cavity of the filters, the refractive index of the sensor material and the cosine of the light incidence angle^56^. The spatial resolution of the sensor is 272 × 512 pixels.

**Figure 7.**
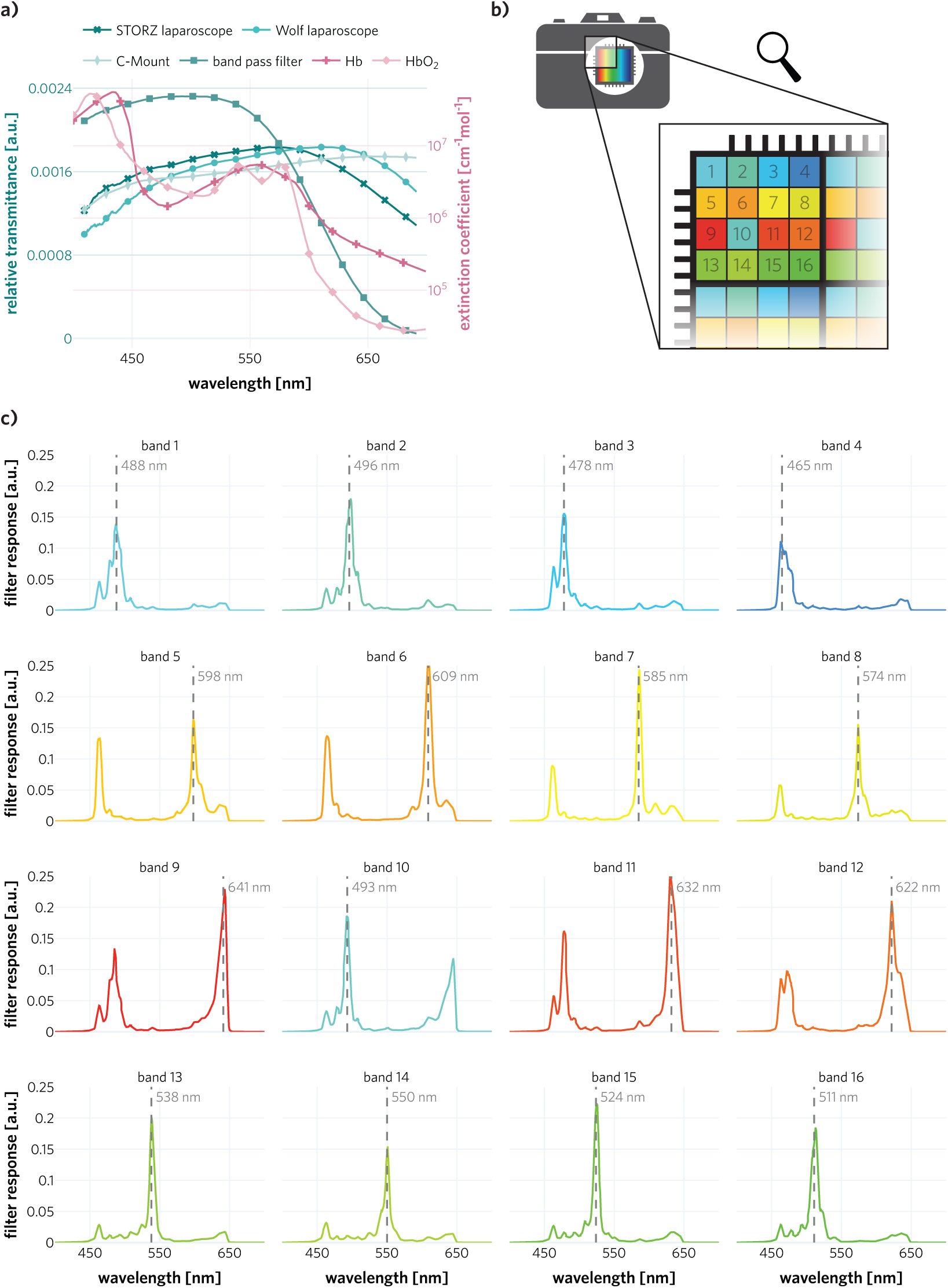
Optical properties of our multispectral system. **a)** *ℓ*_1_ normalized transmission spectra of the laparoscopes, bandpass filter and C-Mount adapter are shown in the left axis. Extinction coefficients of oxyhemoglobin (HbO_2_) and deoxyhemoglobin (Hb) are shown on the right axis. The bandpass filter mainly filters light in the low wavelength region where blood absorbs the most (below ≈ 600 nm), thus making the spectral power distribution that reaches the camera detector more uniform across wavelengths. **b)** Representation of the 4 × 4 mosaic pattern of the multispectral camera sensor. Each colored square represents a different filter; these filters form a 4 × 4 pattern that extends over the whole image. **c)** The filter responses of the multispectral camera bands. Some bands, such as 5–12, show two extra peaks in the spectral response, which are referred to as “second order” peaks.

#### Surgery-specific components

Due to intrinsic optical tissue properties, the reflectance of human tissue in the red region (above ≈ 620 nm) is higher than that in the blue region (below ≈ 490 nm). To ensure more balanced camera counts, and thus similar noise levels across different camera filters, a 335–610 nm bandpass filter (FGB37, Thorlabs Inc., Newton, New Jersey, United States) was placed between the C-Mount adapter and the laparoscope. The C-Mount adapter (20200043, KARL STORZ SE & Co. KG, Tuttlingen, Germany) features an adjustable focal length, with a maximum of 38 mm. To enable the recording of MSI images during minimally invasive surgery, the camera was connected to standard 30° laparoscopes (26003BA, KARL STORZ SE & Co. KG, Tuttlingen, Germany and Panoview, Richard Wolf, Knittlingen, Germany) via the aforementioned C-Mount adapter. The *ℓ*_1_ normalized transmission spectra of the bandpass filter, the laparoscope and the C-Mount adapter are shown in Fig. 7. The extinction coefficients of HbO_2_ and Hb are also shown as a reference^57^. As light source, we chose a Xenon light source (IP20, Richard Wolf GmbH, Knittlingen, Germany) as it provides brighter and more uniformly distributed light across different wavelengths in comparison to a halogen light source.

#### Image recording and storing

For image acquisition and recording, a standard laptop (Msi GE75 Raider 85G, Intel i7, NVIDIA RTX 2080) was used. A custom C++ software (not publicly available) based on the XIMEA Application Program Interface (XiAPI, XIMEA, Münster, Germany) was implemented and used to record the MSI images.

### 5.2 Algorithm for live ischemia monitoring

We phrase the challenge of live ischemia monitoring as an OoD detection problem that only relies on data of the current patient. This mitigates the challenge of acquiring large amounts of annotated training data and the limited robustness which traditional machine and DL approaches suffer from due to high variability of human tissue, acquisition protocols, camera setups, etc. Specifically, our algorithm is trained to compute the likelihood of normal tissue perfusion based on a (several seconds) video sequence acquired at the beginning of each surgery. In the following paragraphs, we review the principle of WAIC^43^, explain how to leverage INNs to compute it in real time and present our novel OoD-based *ischemia index*.

#### WAIC

While we are not aware of any previous work in OoD detection in the field of optical imaging, the topic of OoD has gained increasing interest in the machine learning community. We build our method upon the work by Choi et al.^58^ who proposed WAIC as a means to measure the closeness of a new sample to the training distribution. In Watanabe et al.^43^, WAIC was defined as

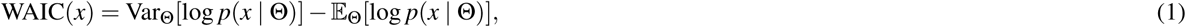

where Θ represents the parameters of the model family that parametrize *p* and the variance and expectation is taken over Θ ∼ *p*(Θ | *X* ^tr^) of the training distribution *X* ^tr^. Then WAIC(*x*) estimates the proximity of a sample *x* to *X* ^tr^. Choi et al.^58^ suggested to use WAIC as a means for OoD in the setting of neural networks. The variance term in equation (1) measures the certainty of the posterior distribution about *p*(· | Θ) for a sample *x*. The intuition behind the term is that it should be more certain about samples the closer the samples are to the training distribution. The expectation term in equation (1) is taken directly from standard density estimation approaches to OoD detection. If the sample *x* lies in a low density region, then it is expected to be OoD. Hence, the log-density is subtracted as another OoD contribution to the WAIC score. A major challenge related to the implementation of WAIC is the efficient computation of log *p*(*x* | Θ). We propose applying INNs to be able to compute log *p*(*x* | Θ) and thus WAIC(*x*) in real time during a surgical procedure.

#### Invertible Neural Networks (INNs) for computing WAIC

Our approach to computing log *p*(*x* | Θ) is based on two steps: (1) leveraging INNs for converting a measurement into a space in which we can analytically compute the log-likelihood and then (2) applying the change of variable formula to obtain log *p*(*x* | Θ). More specifically, we use INNs^59^ based on the *normalizing flow* architecture^60^ to encode the spectra in image space *X* in a latent space *Z* in which the samples are distributed according to a multivariate standard Gaussian. Let *f*_Θ_ : *X* ⊂ ℝ^*n*^ → *Z* ⊂ ℝ^*n*^ denote the neural network with parameters Θ. Then we can use the change of variable formula to compute the log-likelihood log *p*(*x* | Θ) for a spectrum *x* as

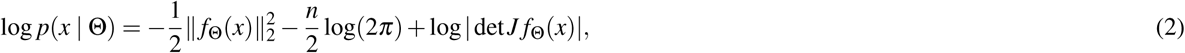

where *J f*_Θ_ denotes the network’s Jacobian and we already assume that the latent space *Z* is normally distributed.

The change of variable formula enables us to compute log *p*(*x* | Θ) for a single neural network. To be able to compute the WAIC variance and expectation term, we generate an ensemble of networks (default: *n* = 5) with identical architecture but different random seeds.

#### Ischemia index

As not only the inference but also the training needs to be performed during the actual surgical procedure, we faced the requirement of fast training times (seconds rather than minutes or hours). To achieve fast network convergence based on the patient-individual data, we pre-trained our neural network ensemble on simulated data. Specifically, we simulated light transport in tissue using a MC method to compute high-resolution spectra covering a large space of physiological states (see Sec. 5.4). In a second step, these high resolution spectra were adapted to the MSI sensor taking the light source, filter responses and other transmission spectra into account. The fully-trained network then allows us to compute WAIC(*x*) for any new spectrum *x*. Higher WAIC values imply that the spectrum is OoD, i.e. ischemic. As the recorded organ of interest consists of more than one multispectral pixel, we post-process the WAIC values by aggregating them over ROIs. This leads to a single value per MSI frame which we refer to as *ischemia index*.

Note that we presented the general idea of using INNs for OoD in the context of uncertainty quantification at a conference workshop^61^. However, the idea of phrasing the ischemia detection problem as an OoD problem is entirely novel (patent pending). The concrete instantiation of this approach for ischemia detection in partial nephrectomy is provided in Sec. 5.4, including all relevant implementation details.

### 5.3 Patient study

The aim of this study was to investigate the feasibility of our approach to contrast agent-free real-time ischemia monitoring. In this section, we describe our data set (see Sec. 5.3.1), the instantiation of our ischemia detection method (see Sec. 5.3.2), and the visualization and evaluation techniques (see Sec. 5.3.3).

#### 5.3.1 Patient data

Here we describe the patient recruitment, image acquisition process, technical inclusion criteria for our evaluation, and our data splits.

##### Patient recruitment

All patients were recruited in the Städtisches Klinikum Karlsruhe (Karlsruhe, Germany). The experiments involving humans were performed in accordance with the Declaration of Helsinki and all protocols were approved by the Landesärztekammer Baden-Württemberg (DE/EKBW01, study reference number: B-F-2019-101). The inclusion criteria for all subjects were: a) subjects were undergoing partial nephrectomy and b) subjects were adults (≥ 18 years old). This resulted in a total of 10 subjects with an age range between 40 and 82 years old, 7 of which were male and 3 female. Tab. 2 shows an overview of all the subjects recruited for this study.

**Table 2.** Patients recruited in our clinical study. **ccRCC:** Clear Cell Renal Cell Carcinoma, **PRCC:** Papillary Renal Cell Carcinoma, **ChRCC:** Chromophobe Renal Cell Carcinoma

| Patient ID | BMI | Sex | Tumor type | Data split |
| --- | --- | --- | --- | --- |
| 1 | 25 | male | PRCC | retrospective |
| 2 | 22 | female | Benign Oncocytoma | retrospective |
| 3 | 40 | male | ccRCC | retrospective |
| 4 | 30 | male | Benign Oncocytoma | retrospective |
| 5 | 49 | female | Leiomyoma | retrospective |
| 6 | 22 | male | ccRCC | prospective |
| 7 | 29 | male | ccRCC | prospective |
| 8 | 43 | male | PRCC | prospective |
| 9 | 31 | male | ChRCC | prospective |
| 10 | 24 | female | Angiomyolipoma | prospective |

##### *In vivo* image acquisition

Recordings were taken from subjects undergoing partial nephrectomy at the Städtisches Klinikum Karlsruhe. The standard procedure involves the generation of six surgical ports, three of which were dedicated to the da Vinci^®^ robotic arms (Intuitive Surgical Deutschland GmbH, Freiburg, Germany), one of which was used for the da Vinci^®^ RGB surgical camera and the remaining two as assisting ports used for other instruments (e.g. scissors, clip appliers, etc.).

The conventional workflow was adapted as follows for our measurements. After removing the fatty tissue from the surface of the kidney and locating the renal artery, the multispectral laparoscope was inserted through one of the assisting ports and three measurements were performed to enable training and validation of our ischemia detection algorithm. Each measurement was performed for 45 s, yielding ≈ 1200 recorded MSI images (see Fig. 8):

**Figure 8.**
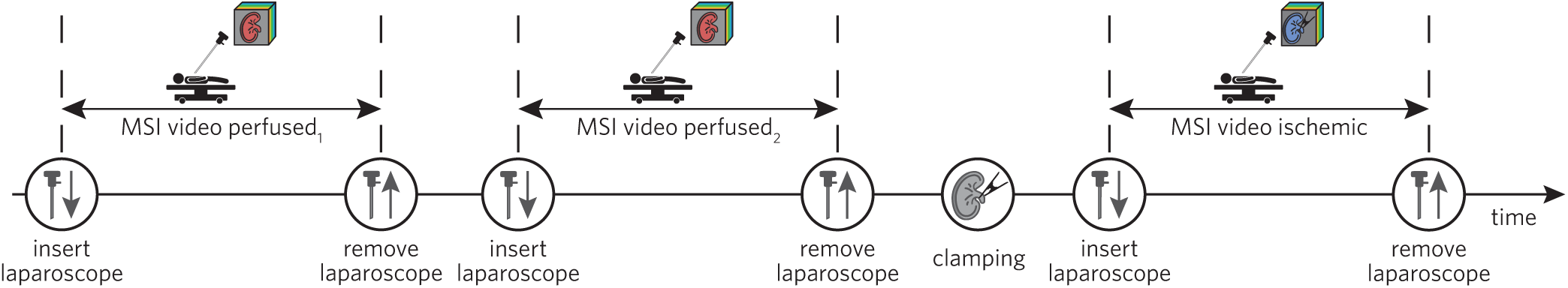
Multispectral imaging (MSI) recording procedure in the operating room (OR). We recorded two MSI sequences (perfused_1_ (for training) and perfused_2_ (for testing)) before the clamp was applied to the renal artery, and one sequence after applying the clamp (for testing). The laparoscope was removed and re-inserted in the patient’s abdominal cavity before recording the testing sequence for perfused kidney to obtain the same conditions as for the ischemic kidney.

1. **First recording of perfused kidney** (training sequence: perfused_1_): MSI images of the surface of the kidney were recorded before a clamp was applied to the renal artery. Part of this data was used for training the personalized *ischemia index*, as detailed in Sec. 5.4.
2. **Second recording of perfused kidney** (testing sequence: perfused_2_): To simulate unsuccessful clamping, an additional image sequence was recorded before the clamp was applied. For this purpose, the multispectral laparoscope was removed from the surgical port and re-inserted through the same port. Then, MSI images of the surface of the kidney were again recorded. The re-insertion of the laparoscope was performed to obtain slightly different acquisition conditions. This is also the case when actual clamping is performed, as the laparoscope needs to be removed before the renal artery can be clamped through the same surgical port (see next paragraph).
3. **Recording of ischemic kidney** (testing sequence: ischemic): The laparoscope was removed from the abdominal cavity, such that the surgical team could perform the clamping procedure through the same port used for the laparoscope. The renal artery was then clamped, the laparoscope was re-inserted in the abdominal cavity for MSI acquisition. Afterwards, the success of the clamping procedure was confirmed by ICG injection into the subject’s bloodstream and subsequent visualization of the fluorescence signal with the Firefly system of the da Vinci^®^ robot. The ICG injection was prepared by mixing 50 mg of ICG powder (PULSION Medical Systems SE, Feldkirchen, Germany) with 10 ml of distilled water.

To obtain high-quality data, all measurements were performed on parts of the tissue in which regions without blood and fatty tissue could be observed. Prior to each measurement, a) the camera integration time was set to 40 ms, b) the laparoscope was positioned to include the clean surface of the kidney in the field of view of the camera and c) the intensity of the light source was adjusted to minimize the number of specular reflections observed on the surface of the kidney. Furthermore, the light source of the da Vinci^®^ RGB surgical camera was turned off during all our measurements.

##### Study cohort

To obtain a reliable reference for our measurements, we only included patients in which the ischemia induction of the kidney was successfully confirmed by ICG injection as part of the traditional surgical procedure. Furthermore, we omitted patients for which image recordings were underexposed due to a faulty light guide. A common flaw in machine learning-based image analysis is the overfitting on the test data^62^. Even if an algorithm is strictly trained on the training data, a common procedure involves testing multiple different models or hyperparameter configurations on the test set and then reporting the best model. To avoid an overestimation of algorithm performance resulting from such practice, we finalized the complete algorithm based on existing data sets before conducting the patient study. The only exception was the number of MSI frames used for training the networks used to compute the *ischemia index*. To fix this hyperparameter, we separated the recruited patients into a retrospective and prospective split. The retrospective data split was composed of the first half of the patients (5) and the prospective split of the second half of the patients (5). The number of MSI frames was determined using only the retrospective data set.

#### 5.3.2 Automatic ischemia detection

To instantiate our ischemia monitoring approach (see Sec. 5.2) for partial nephrectomy, the following design decisions needed to be made: (1) How to preprocess the raw camera data before providing it to the neural network ensemble? (2) How to aggregate values over individual spectra in order to derive an image-level *ischemia index* at test time?

##### Data preprocessing

We decided to represent the tissue state in an MSI image by two ROIs that correspond to regions on the tissue from which high-quality measurements can be obtained. The ROIs were chosen by the physician or technical assistant at the beginning of a recording sequence according to the process detailed in Sec. 5.4. As we implemented an automatic ROI tracking algorithm, the ROIs only had to be chosen once at the beginning of each sequence. The data from each ROI was first normalized with a white (*W*) and dark (*D*) reference recording taken with a highly reflective target (Spectralon^®^, Edmund Optics, Barrington, USA). Given an ROI of dimensions *N* × *M* × *B*, where (*N, M*) are the spatial dimensions and *B* is the number of spectral bands, the intensity *I* of each pixel at spatial location (*i, j*) and spectral band *k* was normalized according to:

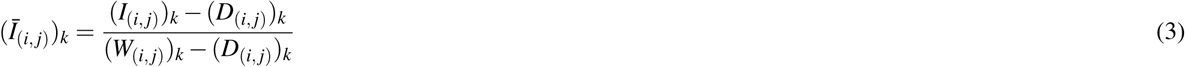

Subsequently, an *ℓ*_2_ normalization across different bands was performed to compensate for the influence of light source intensity changes due to changes in the distance of the laparoscope to the surface of the kidney. The resulting spectra (*Î*_(*i, j*)_)_*k*_ can be compared between different image sequences. The normalized spectra *Î* were then used to train our ensemble of INNs, and the median normalized spectra within each ROI were used for the rest of our analysis.

##### Ischemia index

Our ischemia monitoring approach leveraged spectral information from two different ROIs. To compute our *ischemia index* from this information, we first aggregated WAIC values belonging to the same ROI,

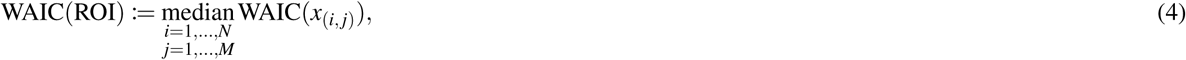

where (*i, j*) is taken over the spatial dimensions of the ROI of shape *N* × *M*.

Then, we aggregated the two ROIs per frame via the mean to obtain the final *ischemia index*

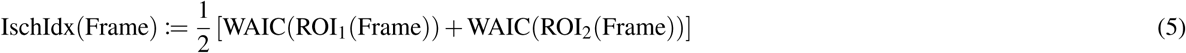

For ease of visualization, we min-max-normalized the *ischemia index* for each patient individually. As this is a strictly monotonic transformation, this has no influence on the AU-ROC metric.

#### 5.3.3 Assessment methods

This section introduces our methods for assessing tissue heterogeneity and for validating our *ischemia index*.

##### Statistical analysis of tissue heterogeneity

For 2D visualization of the high-dimensional spectral data, the data from each ROI was first normalized according to the procedure described in Sec. 5.3.2 yielding a median spectrum per ROI, frame, state and subject. For each frame, we averaged the median spectra across all ROIs, computed the first two PCs based on the data from all subjects and projected the data points onto these new axes^42^.

We further computed the proportion of explained variability in reflectance by different components using linear mixed models, as suggested by Schreck et al.^63^. Given a patient index *i* = 1, …, 10, and an MSI frame index *j* = 1, …, *n*_*i*_, we fitted the following model for each wavelength separately:

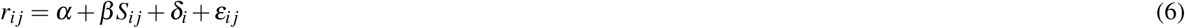

where *r* represents the averaged median reflectance across tracked ROIs for a given wavelength (see Sec. 5.4). *S*_*i j*_ is an indicator variable, indicating the perfusion state of the kidney (1 for ischemic or 0 for perfused). Furthermore, *α* represents an intercept for each linear model, *β* is a fixed state effect, 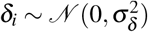 is a random patient effect and 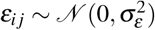 are residuals, with respective variances 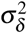 and 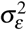 that are assumed to be independently normally distributed. We computed the proportions of explained variability following^63^.

##### Validation of ischemia index

In order to validate the *ischemia index*, we designed the recording process such that we had comparable testing sequences for perfused and ischemic kidney as detailed in Sec. 5.3.1. For performance assessment, we computed the *ischemia index* for the first 70 frames of both testing sequences. Using ischemic as the positive class, perfused as the negative class, and the *ischemia index* as prediction score (higher implying more ischemic), we then computed the AU-ROC as primary performance metric.

##### Analysis of example patients

For the detailed analysis of the representative patient (patient 3) and the patient on which our method failed (patient 7; Fig. 6), we computed the box plots on the same data as our network was trained and tested on, that is, all valid pixels from three kidney states (perfused_1_, perfused_2_ and ischemic), both ROIs, and the first 70 frames were used. The same data was also used for the density plots where we computed the first two PCs and then applied KDE estimation^64, 65^ on the projected values for each state separately via the fast KDE method^66, 67^.

### 5.4 Implementation details

This section presents implementation details for some of the image analysis methods presented in the manuscript.

#### INNs

The training data consisted of two ROIs of size 30 × 30 pixels located on the perfused kidney. These ROIs were tracked for 70 consecutive frames. The INNs were implemented using the PyTorch framework and the FrEIA package for invertible architectures. Following up on our previous work with INNs^59, 61, 68^, we applied the *normalizing flow* architecture originally introduced in Dinh et al.^60^ and used the following network default settings: 20 affine coupling blocks^60^ with 3 layer fully-connected subnetworks with 256 hidden dimensions, rectified linear unit (ReLU) activations and fixed channel-permutations. The networks were trained using Maximum-Likelihood training, i.e. by minimizing the loss *L*(*x*) = −log *p*(*x* | Θ) as given in Equation (2) using the Adam optimizer^69^, a learning rate of 1 · 10^−4^ and weight decay of 1 · 10^−4^. The training data was z-score normalized and we used noise augmentation using additive Gaussian noise with a standard deviation of 0.05. The pre-training based on MC simulations was performed offline with 100 epochs. Fine-tuning on the patient data was done for 10 epochs. All hyperparameters, except for the number of training frames, were optimized with pre-existing data disjoint from the current study data.

#### ROI tracking

At least two ROIs of size 30 × 30 px were selected on each image sequence, each based on the requirements that a) there was no adipose tissue on top of the kidney tissue, b) there was no blood leakage staining the kidney tissue, c) the camera counts were not oversaturated and not undersaturated, d) there was no visible smoke, and e) the ROI locations did not overlap. Each ROI was defined in the first image and subsequently tracked across consecutive frames with a deep learning-based algorithm. As the multispectral laparoscope needed to be retracted between the two recordings (before and during clamping), we were not able to ensure imaging of the exact same region. Hence, the ROIs of the different sequences do not correspond.

The tracking was performed on RGB images reconstructed from the MSI images, as detailed in Sec. 5.4. In order to enable reliable tracking, the RGB images were first transformed to HSV (hue, saturation value) color space, then the V channel was normalized with Contrast Limited Adaptive Histogram Equalization (CLAHE), and finally the resulting image was converted back to RGB. The kernel size used by CLAHE was set to 1/8 of the image height by 1/8 of its width, the number of bins was set to 256 and the clipping limit to 0.01.

The reconstructed and normalized RGB images were fed into a pre-trained VGG19^70^ neural network and deep features were extracted from its seventh convolutional layer. The extracted features were further processed by the tracker Discriminative Correlation Filter with Channel and Spatial Reliability (CSR-DCF)^71^. This tracker compares the features extracted from two consecutive frames in order to localize the ROI in the new frame.

Before using a processed video sequence (70 frames corresponding to the 2–3 s) for training or testing, the sequence of automatically tracked ROIs was manually verified to ensure that a) the ROIs did not disappear from the camera field of view, b) at least 95 % of all tracked pixels were not oversaturated and not undersaturated, and c) there was no visible drift of the ROI from the original annotated tissue region. Those two ROIs fulfilling the criteria that could be tracked the longest were kept for further processing.

#### Monte Carlo simulations

Generation of simulated tissue spectra for pretraining the *ischemia index* was inspired by previous work^12^. Light transport of several tissue structures was simulated with a model composed of three infinitely wide slabs. Each slab was defined by optical and physiological parameter values of blood volume fraction *v*_hb_, blood oxygen saturation *s*, reduced scattering coefficient at 500 nm *a*_mie_, scattering power *b*_mie_, anisotropy *g*, refractive index *n* and layer thickness *d*. Such parameters were computed based on reference values from literature^72^ and later used to create MC simulations. The reference values used from literature include: extinction coefficients of deoxyhemoglobin *ε*_Hb_ and oxyhemoglobin *ε*_HbO2_, absorption *µ*_*a*_ and scattering *µ*_*s*_ coefficients. A Graphics Processing Unit (GPU) accelerated version^73^ of the popular Monte Carlo Multi-Layered (MCML) simulation framework^74^ was chosen to generate spectral reflectances. The ranges from which the parameters were uniformly sampled as well as general simulation parameters are summarized in Table 3.

**Table 3.** The simulated ranges of physiological parameters, and their usage in the simulation setup. Here *v*_Hb_ represents the blood volume fraction, *s* the blood oxygen saturation, *a*_*mie*_ the reduced scattering coefficient at 500 nm, *b*_*mie*_ the scattering power, *g* the tissue anisotropy, *n* the refractive index and *d* the tissue thickness and *ε* the extinction coefficients of oxyhemoglobin (HbO_2_) and deoxyhemoglobin (HbO). All parameters were uniformly sampled within the specified range.

| | $v_{\text{Hb}}[\%]$ | $s[\%]$ | $a_{\text{mie}}[\text{cm}^{-1}]$ | $b_{\text{mie}}[\text{a.u.}]$ | $g[\text{a.u.}]$ | $n[\text{a.u.}]$ | $d[\text{cm}]$ |
| --- | --- | --- | --- | --- | --- | --- | --- |
| layer 1: | 0–30 | 0–100 | 5–50 | 0.3–3 | 0.80–0.95 | 1.33–1.54 | 0.002–0.2 |
| layer 2: | 0–30 | 0–100 | 5–50 | 0.3–3 | 0.80–0.95 | 1.33–1.54 | 0.002–0.2 |
| layer 3: | 0–30 | 0–100 | 5–50 | 0.3–3 | 0.80–0.95 | 1.33–1.54 | 0.002–0.2 |
| $\mu_a(v_{\text{Hb}}, s, \lambda) = v_{\text{Hb}}(s \cdot \epsilon_{\text{HbO}_2}(\lambda) + (1 - s) \cdot \epsilon_{\text{Hb}}(\lambda)) \cdot \ln(10) \cdot 150 \text{g L}^{-1} \cdot (6.45 \times 10^4 \text{g mol}^{-1})^{-1}$ | | | | | | | |
| $\mu_s(a_{\text{mie}}, b, \lambda) = \frac{a_{\text{mie}}}{1-g} \left( \frac{\lambda}{500 \text{nm}} \right)^{-b_{\text{mie}}}$ | | | | | | | |
| simulation framework: GPU-MCML<sup>73</sup>, $10^6$ photons per simulation | | | | | | | |
| simulated samples: $5.5 \times 10^5$ | | | | | | | |
| sample wavelength range $[\lambda_{\min} - \lambda_{\max}]$: 300–1000 nm, step size 2 nm | | | | | | | |

The simulated reflectances *r*(*λ*) were then transformed into the MSI camera measurement *r*_*k*_ at band *k* according to:

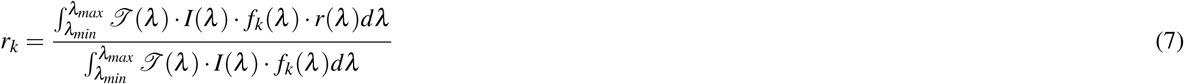

Here, *ℐ*(*λ*) represents the optical transmission profile of the optical components of our hardware setup (see Fig. 7), *I*(*λ*) is the relative irradiance of the light source, and *f*_*k*_(*λ*) characterizes the *k*th optical filter response of the camera. The transformed simulated spectra *r*_*k*_ were used to pre-train our DL models as described in Sec. 5.3.2.

#### RGB image reconstruction

Reconstruction of RGB images from MSI was required for (1) automatic ROI tracking and (2) illustration purposes. The reconstruction was achieved by computing a transformation matrix *T* of dimensions (3 × *B*), where *B* = 16 represents the number of bands in one MSI image. A linear regressor was fitted to compute the transformation matrix *T* based on the filter response of the MSI camera, the transmission of each optical component (see Fig. 7) and the filter response of an artificial RGB camera. The artificial RGB camera was simulated by 3 Gaussian filters centered at 460 nm, 550 nm and 640 nm, each with a standard deviation of 42 nm. In addition, the spectral response of the MSI camera was adjusted by multiplying the transmission ℐ of each optical component by the filter response of the MSI camera *F*_*MSI*_.

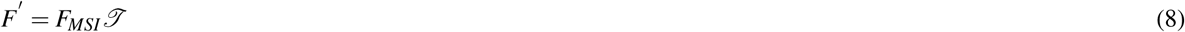

If we consider the filter response of the artificial camera *F*_*RGB*_ and the spectral response of the MSI camera *F′* after correcting for the optical components integrated in the system, we can compute the transformation matrix as:

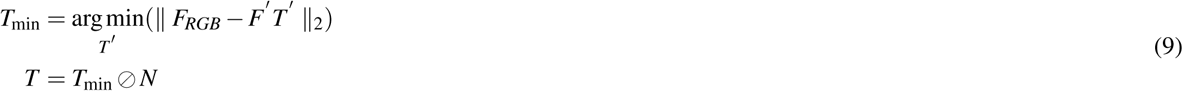

where *T* represents the coefficients of the linear regressor, ⊘ denotes component-wise division, and *N* is a normalization vector with 3 elements, which can be computed as:

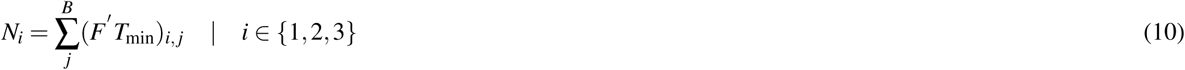

#### Optical transmission profiles

All optical transmission profiles shown in Fig. 7 a) were measured with a spectrometer (HR2000+, Ocean Insight, Orlando, USA). 100 transmission measurements of each optical component were averaged and then smoothed across wavelengths by rolling window averaging with a window of ≈ 19 nm width. The values in the ranges 400–419 nm and 681–700 nm were ignored after smoothing in order to avoid unwanted border effects.

#### Beer-Lambert regression

We used the Beer-Lambert law^49^ to estimate the total blood volume fraction (*v*_HbT_) and oxygenation (*s*) within the tracked ROIs described in Sec. 5.4. This law states that a linear relationship exists between the absorption *a*(*λ*) of a media, its attenuation coefficient *µ*_*a*_(*λ*) and the optical path length *l*:

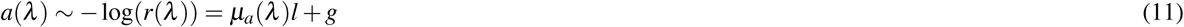

where *g* is a term accounting for scattering losses. Taking into account that blood is the main absorber in internal organs, we can replace the attenuation coefficients by the extinction coefficients of blood

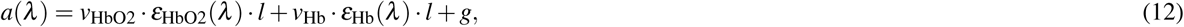

where *v* are concentrations and *ε* the corresponding extinction coefficients. Given measurements at multiple wavelengths, equation (12) describes a system of linear equations, which can be solved for *v*_HbO2_ · *l* and *v*_Hb_ · *l* using ordinary least squares regression. This enables us to estimate the oxygenation *s* and the total hemoglobin concentration *v*_HbT_ via:

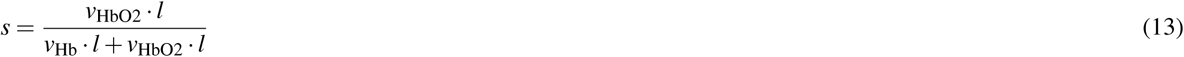

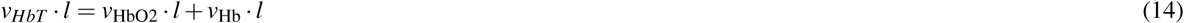

Note that estimations of *v*_*HbT*_ can only be computed up to a constant factor *l*. Since all extinction coefficients are given at a high spectral resolution, but we use a multispectral sensor with a relatively low number of bands, we adapted the high resolution extinction coefficients by averaging them over the filter response function of each band.

## Supporting information

Supplemnetal ROI tracking diagram

## Data Availability

The raw in vivo data used in this manuscript is not available because it was not indicated in the patients' signed consent.

## Data availability

The raw *in vivo* data used in this manuscript is not available because it was not indicated in the patients’ signed consent.

## Code availability

The source code of our core methodology cannot be open-sourced at the moment, because a patent involving this method is under review.

## Acknowledgements

This project has received funding from the European Research Council (ERC) under the European Union’s Horizon 2020 research and innovation programme (NEURAL SPICING, Grant agreement No. 101002198), and the ERC starting Grant COMBIOSCOPY under Grant agreement No. ERC-2015-StG-37960. This project also received support from the Helmholtz Association under the joint research school “HIDSS4Health – Helmholtz Information and Data Science School for Health”. Part of this work was funded by the Helmholtz Imaging Platform (HIP), a platform of the Helmholtz Incubator on Information and Data Science. Some icons showed in the figures were provided by Freepik from Flaticon.com.

## Ethics declaration

All experiments involving humans were performed in accordance with the Declaration of Helsinki and all protocols were approved by the Landesärztekammer Baden-Wuerttemberg (DE/EKBW01, study reference number: B-F-2019-101). The study is registered with the German Clinical Trials Register (DRKS00020996).

