## Supplementary figures and images for "Spectral imaging enables contrast agent-free real-time ischemia monitoring in laparoscopic surgery"

### Supplemnetal ROI tracking diagram

#### Region of interest annotation diagram

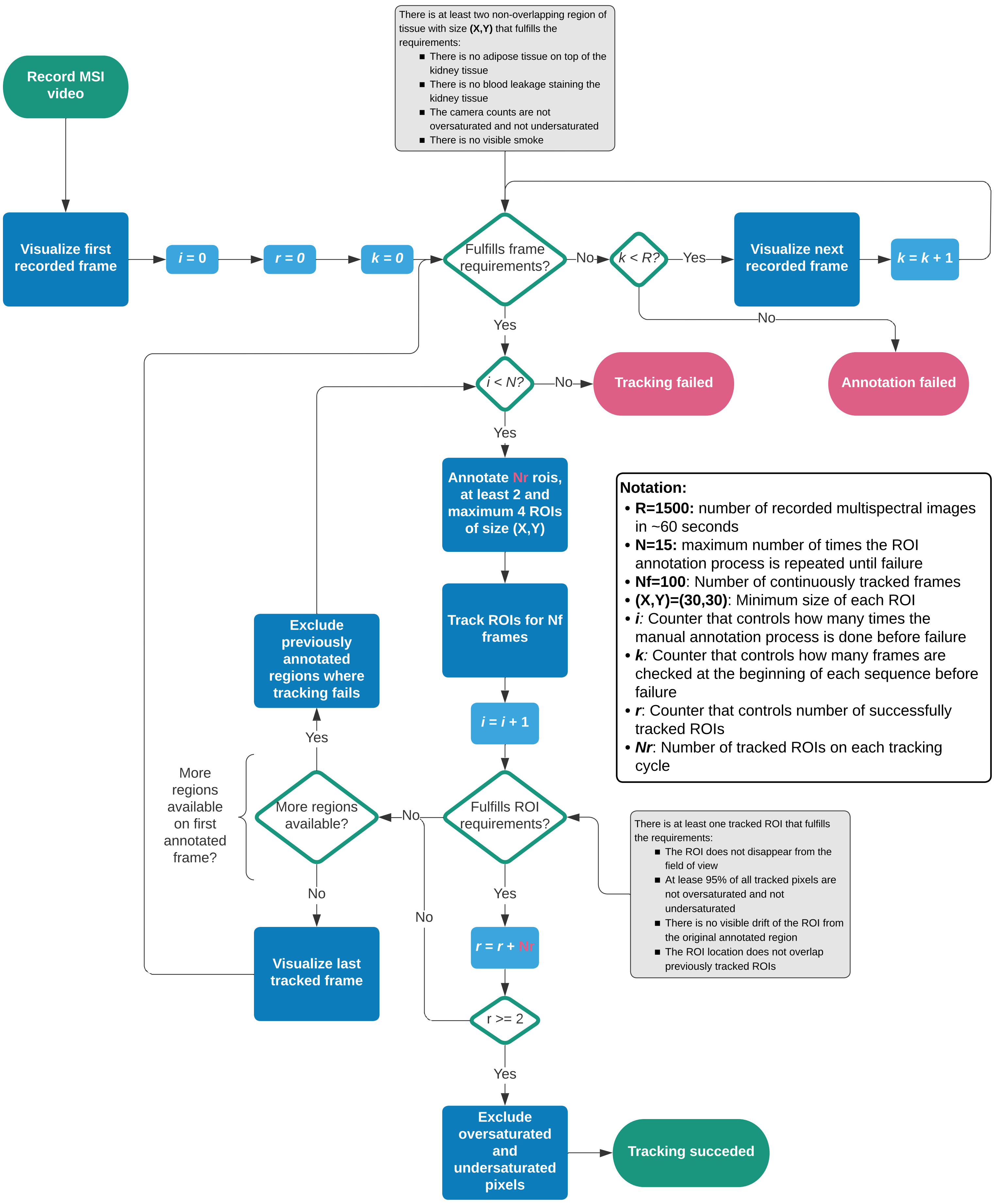
